# Recipient Cells Are the Source of Hematologic Malignancies After Graft Failure and Mixed Chimerism in Adults with SCD

**DOI:** 10.1101/2025.01.17.25320670

**Authors:** Mohamed A. E. Ali, Emily M. Limerick, Matthew M. Hsieh, Kalpana Upadhyaya, Xin Xu, Oswald Phang, Jean Pierre Kambala Mukendi, Katherine R. Calvo, Maria Lopez-Ocasio, Pradeep Dagur, Courtney D. Fitzhugh

## Abstract

Non-myeloablative hematopoietic cell transplantation (HCT) is a curative option for individuals with sickle cell disease (SCD). Our traditional goal with this approach has been to achieve a state of mixed donor/recipient chimerism. Recently, we reported an increased risk of hematologic malignancies (HMs) in adults with SCD following graft failure or mixed chimerism. To evaluate the origin of HMs, we performed chimerism analyses of 5 patients with SCD who developed HMs after non-myeloablative HCT. DNA was extracted from sorted peripheral blood or bone marrow cells representing mature cell lineages or leukemic blasts and subjected to chimerism analysis by PCR amplification of polymorphic short tandem repeats. Unlike mature cell lineages in patients with mixed chimerism, which still showed a donor-derived fraction of cells, leukemic blast cells were found to be 99-100% recipient-derived in all patients. Non-myeloablative conditioning allows for the survival of patients’ cells that might harbor pre-leukemic clones that possess the capacity to evolve under genotoxic or environmental stress into malignancies; therefore, we have modified our HCT protocols with the goal of full donor chimerism to mitigate the risk of HM development.

## Introduction

Hematopoietic cell transplant (HCT) is the only curative option for individuals with sickle cell disease (SCD). Traditionally, young patients with a human leukocyte antigen (HLA)-matched donor received myeloablative conditioning followed by HCT to completely replace recipient hematopoietic cells with healthy donor cells (full donor chimerism). Unfortunately, less than 15% of patients have an HLA-matched sibling donor (MSD). Conversely, haploidentical (haplo) HCT greatly expands the donor pool. Moreover, adults with SCD and overt organ damage cannot tolerate myeloablative conditioning. Therefore, we developed a non-myeloablative approach that could be applied to such patients. This approach initially aimed at achieving a state of mixed donor and recipient chimerism, where the hematopoietic system is constituted by both the donor’s and recipient’s cells. Indeed, we have reported that 20% donor myeloid chimerism is sufficient to reverse the SCD phenotype because of a significant difference in the life span of healthy versus sickled red blood cells ^1^.

In 2023, the FDA approved two additional potentially curative therapies. Firstly, Lyfgenia, by Bluebird Bio, uses a lentiviral vector to genetically modify hematopoietic stem and progenitor cells (HSPCs) to produce HbA^T87Q^, which functions similarly to HbA. Secondly, Casgevy, a clustered, regularly interspaced palindromic repeats-associated protein-9 nuclease (CRISPR-Cas9) gene-edited therapy, reactivates the production of fetal hemoglobin. Both therapies would still require myeloablative conditioning, which limits its application to younger patients with no significant organ damage.

Two studies have reported an increased relative risk of leukemia development in patients with SCD, though the absolute risk is low ^2^. Moreover, several groups have reported an increased risk of leukemia development after curative therapies, mainly after graft failure in SCD patients ^2^. We have recently reported that eight out of 120 patients who received non-myeloablative allogeneic HCT developed hematologic malignancies (HMs) between 4 months and 9 years post-HCT: five developed aggressive therapy-related myeloid neoplasms (myelodysplastic syndrome (MDS) or acute myeloid leukemia (AML)), one T-cell acute lymphoblastic leukemia (ALL), one chronic myeloid leukemia, and one mantle cell lymphoma ^2^. In addition, two adult Group A patients from the bluebird bio study subsequently developed AML ^2^.

Leukemia development has been associated with the acquisition of somatic mutations, which accumulate with aging in a condition known as clonal hematopoiesis (CH) ^3^. Somatic mutations in genes involved in stem cell self-renewal or cellular differentiation pathways bestow a considerable survival advantage or fitness upon the mutant hemopoietic clone, resulting in its significant expansion compared to non-mutant cells ^4^. SCD has been reported to be significantly associated with increased prevalence of CH; among the most commonly mutated genes is *TP53*, which comes in second, accounting for 13% of all CH mutations in SCD ^4^. CH mutations have been reported in patients with SCD who developed HMs after curative therapies ^4,5^. The origin of the HMs, donor- or recipient-derived, has not been previously reported. Since most of those HMs developed after graft failure, we hypothesized that the origin of the leukemia-initiating cells (LICs) is the recipient due to a survival advantage of HSPCs containing somatic mutations that transform with genotoxic conditioning followed by erythropoietic stress. In this report, we investigated the source of leukemic blasts and MDS mononuclear cells (MNCs) by analyzing the chimerism levels in peripheral blood (PB) or bone marrow (BM) cells from patients who developed HMs post-HCT.

## Materials and Methods

We utilized frozen biospecimens from patients diagnosed with the most aggressive HMs after HCT. All samples were collected under an NHLBI IRB-approved protocol, clinicaltrials.gov identifiers NCT00977691, NCT00061568, and NCT02105766. SCD-03 and SCD-04 MNCs were stained using Biolegend’s FITC anti-CD45 (Cat# 304006), BV785 anti-CD3+ (Cat# 317329), BV711 anti-CD19 (Cat# 302245), PE anti-CD33 (Cat# 366607), PE-Cy7 anti-CD34 (Cat# 343515), and, BV421 anti-CD13 (Cat# 301715) then, sorted in 4-way purity mode using BD FACSymphony™ S6 Cell Sorter. SCD-05 MNCs were stained using Biolegend’s FITC anti-CD45, BV711 anti-CD19, BV605 anti-CD11b (Cat# 301335), APC anti-CD1a (Cat# 344907), PE-Cy7 anti-CD2 (Cat# 300221), BV421 anti-CD5 (Cat# 364029), PE anti-CD7 (Cat# 395603) and, APC-Cy7 anti-CD38 (Cat# 356615), then sorted in 4-way purity mode using BD FACSAria™ Fusion Flow Cytometer. 7-amino-actinomycin D (7-AAD) (Cat# 130-111-568, Miltenyi Biotec) was used as a viability stain. DNA was extracted from unsorted or sorted PB or BM MNCs using Qiagen DNeasy Blood & Tissue Kit (Cat# 69504) according to the manufacturer’s protocol.

Purified DNA from various cell types was quantified using Nanodrop2000. DNA samples were diluted to 0.5 ng/ul and used for short tandem repeat (STR) loci amplification using the GenePrint®24 system. Amplified PCR products were mixed with HiDi formamide + Internal Lane Standard (WEN ILS600) and heat-denatured on a PCR thermocycler. The denatured products were run on ABI’s 3500 Genetic analyzer for capillary electrophoresis. Raw data from the genetic analyzer was analyzed for peak calling using NCBI’s Osiris software. Peak-called data from Osiris were then imported to FileMaker software, where the STR allele peaks obtained from the sample were compared with those from the patient and donor STR profile previously established in the system. Donor chimerism % calculation was done using selected informative loci in FileMaker software.

## Results and discussion

Five individuals with homozygous SCD (HbSS) were included in this study (Supplementary Table 1): three received MSD, and 2 received haplo HCT with alemtuzumab, 300-400cGy radiation, and sirolimus. Two patients (SCD-01 and SCD-02) received 100mg/kg post-transplant cyclophosphamide (PT-Cy), and two patients (SCD-04 and SCD-05) pentostatin and oral Cy preconditioning. All but one (SCD-05) are now deceased. Two patients, aged 35-40 years at HCT, had MDS 2 and 2.5 years post-HCT (SCD-01 and SCD-02, respectively); one patient, aged 20-25 years at HCT, had AML 4 months post-HCT (SCD-03); a second patient, aged 30-35 years at HCT, had AML 5.5 years post-HCT (SCD-04); and one patient, aged 35-40 years at HCT, had T-cell ALL 3 years post-HCT (SCD-05). Three patients (SCD-01, SCD-02, and SCD-03) had graft failure with 0% PB donor myeloid chimerism (DMC) and 0% donor lymphoid chimerism (DLC) accompanied by the return of SCD. SCD-04 had impending graft failure with 16% PB DMC and 18% DLC. SCD-05 had mixed chimerism with 30% PB DMC and 25% DLC (Table 1).

**Table 1.** Chimerism analysis of sorted cells.

| Patient No. | % PB DMC at dx | % PB DLC at dx | Sorted cell tissue | Sorted cell phenotype | Sorted cell chimerism |  |
| --- | --- | --- | --- | --- | --- | --- |
|  |  |  |  |  | % donor | % recipient |
| SCD-01 | 0 | 0 | BM | Unsorted BM MNCs | 0 | 100 |
| SCD-02 | 0 | 0 | PB | Unsorted PB MNCs | 0 | 100 |
| SCD-03 | 0 | 0 | PB | CD3<sup>+</sup> | 0 | 100 |
|  |  |  |  | CD19<sup>+</sup> | 0 | 100 |
|  |  |  |  | CD3<sup>-</sup>CD19<sup>-</sup>CD13<sup>+</sup>CD33<sup>+</sup>CD34<sup>+</sup> | 0 | 100 |
| SCD-04 | 16 | 18 | PB | CD3<sup>+</sup> | 16 | 84 |
|  |  |  |  | CD3<sup>-</sup>CD19<sup>-</sup>CD13<sup>+</sup>CD33<sup>+</sup>CD34<sup>+</sup> | 1 | 99 |
| SCD-05 | 30 | 25 | BM | CD11b<sup>+</sup> | 11 | 89 |
|  |  |  |  | CD19<sup>+</sup> | 22 | 78 |
|  |  |  |  | CD19<sup>-</sup>CD11b<sup>-</sup>CD2<sup>+</sup>CD5<sup>+</sup>CD7<sup>+</sup>CD38<sup>+</sup>CD1a<sup>-</sup> | 0 | 100 |

Although there were no samples for SCD-01, a whole BM chimerism analysis was performed at the diagnostic evaluation, revealing 100% recipient chimerism. Further, we confirmed the origin of MDS MNCs from SCD-02 by analyzing the chimerism of DNA extracted from PB MNCs at year 3 post-HCT; cells were 100% recipient-derived. Moreover, from a PB sample of SCD-03 at year 5 post-HCT, we sorted 3 PB MNC populations: CD3^+^ T cells and CD19^+^ B cells, representing mature cell lineages, and CD3^-^CD19^-^CD13^+^CD33^+^CD34^+^, representing leukemic blasts (Supplementary Figure 1A). All showed a 100% recipient-derived origin, which is expected since this patient experienced graft failure 90 days post-HCT (Supplementary Table 1). In addition, we sorted two populations from SCD-04 PB at day 100 post-HCT: CD3^+^ T cells and CD3^-^CD19^-^CD13^+^CD33^+^CD34^+^ leukemic blasts (Supplementary Figure 1B and 1C). While we detected 16% donor-derived cells in the T cell compartment, consistent with the PB DLC performed at diagnosis, the blast population showed 99% recipient-derived cells (Table 1). Lastly, we sorted three populations from SCD-05 BM at year 3 post-HCT: CD19^+^ B cells, CD11b^+^ Myeloid cells, and CD19^-^CD11b^-^CD2^+^CD5^+^CD7^+^CD38^+^ CD1a^-^, mainly leukemic blasts (Supplementary Figure 1D and 1E). While we detected 11% BM DMC and 22% BM B-cell chimerism, the blast population was 100% recipient-derived.

In conclusion, we showed that, in patients who experienced MDS, AML, or T-cell ALL following graft failure or mixed chimerism, leukemic blasts or MDS MNCs originated from patient cells, with 99-100% recipient chimerism. Given that CH has been reported to develop in SCD patients at a younger age and that mutations in *TP53*, a major tumor suppressor gene, come in second in prevalence, a possible explanation is the existence of premalignant clones of small sizes within the patients’ HSPCs ^4^. Jones and DeBaun hypothesized that after gene therapy for SCD, the stress of switching from homeostatic to regenerative hematopoiesis by transplanted cells drives clonal expansion and leukemogenic transformation of preexisting premalignant clones, eventually resulting in AML/MDS ^6^. Since we found that the leukemic blasts or MDS MNCs containing the somatic mutations had the survival advantage, being 99-100% recipient-derived, while the more differentiated cells had mixed donor/recipient chimerism, our data support their hypothesis. These clones are exposed to various insults, including HCT conditioning (radiation, chemotherapy, or both), erythropoietic stress as a result of graft failure or gene therapy, especially when the cell dose is suboptimal, as well as alloreactivity in the allogeneic setting, all of which may drive the expansion of clones. Indeed, we have previously reported the discovery of baseline low-level *TP53* mutations that progressively expanded over time until HM diagnosis ^5^. Research is ongoing to study the prevalence of CH in individuals with SCD at baseline and the evolution of CH following allogeneic HCT and gene therapy for SCD to identify genetic risk factors for HM development. Intending to eliminate any premalignant clones, our future protocols aim to achieve full donor chimerism to mitigate the risk of evolving preexisting leukemic clones or transforming recipient, clonally expanded hematopoietic cells.

## Data Availability

All data produced in the present work are contained in the manuscript

## Acknowledgments

This research was supported by the Intramural Research Program of the National Heart, Lung, and Blood Institute (NHLBI), National Institutes of Health and the Cooperative Study of Late Effects for SCD Curative Therapies (COALESCE, 1U01HL156620-01, NHLBI).

## Authorship Contributions

M.A. designed the study, performed the experiments, analyzed the data, and wrote the manuscript. E.M.L. and M.M.H. analyzed the data and reviewed the manuscript. K.U. performed the experiments, analyzed the data, and reviewed the manuscript. X.X, O.P., JP.K.M., K.R.C., M.L.O, and P.D., analyzed the data and reviewed the manuscript. C.F. conceived the study, designed the experiments, analyzed the data, and wrote the manuscript.

## Disclosure of Conflicts of Interest

The authors declare no conflicts of interest.

---

**Supplementary Table 1.** Clinical characteristics of patients who developed hematologic malignancies after HCT for SCD.

| Patient No. | SCD Type | Age at HCT | Sex | HCT Type | TBI Dose cGy | PT-Cy dose mg/kg | Day of Graft Failure | Dx | Time to Dx post-HCT (yr) | Cytogenetics and BM blasts at Dx | Current status |
| --- | --- | --- | --- | --- | --- | --- | --- | --- | --- | --- | --- |
| SCD-01 | HbSS | 35-40 | Male | HLA-matched | 300 | 0 | 183 | MDS | 2.5 | Complex, <5% | Dec |
| SCD-02 | HbSS | 35-40 | Male | Haplo | 400 | 100 | 73 | MDS | 2 | Complex, <5% | Dec |
| SCD-03 | HbSS | 20-25 | Female | Haplo | 400 | 100 | 90 | AML | 5.5 | Complex, 20% | Dec |
| SCD-04 | HbSS | 30-35 | Male | HLA-matched | 300 | 0 | 74 | AML | 0.33 | Complex, 15%-20% | Dec |
| SCD-05 | HbSS | 35-40 | Female | HLA-matched | 300 | 0 | N/A | T-cell ALL | 3 | 46XX, t(9:22)[18]/46,XY[2] BCR/ABL1 p190 fusion, 93% | Alive |

HCT, hematopoietic cell transplant; SCD, sickle cell disease; HbSS, homozygous SCD; TBI, total body irradiation; PT-Cy, post-transplant cyclophosphamide; Dx, diagnosis; Haplo, haploidentical; MDS, myelodysplastic syndrome; AML, acute myeloid leukemia; ALL, acute lymphoblastic leukemia; t, translocation; Dec, deceased; N/A, not applicable.

**Supplementary Figure 1.**
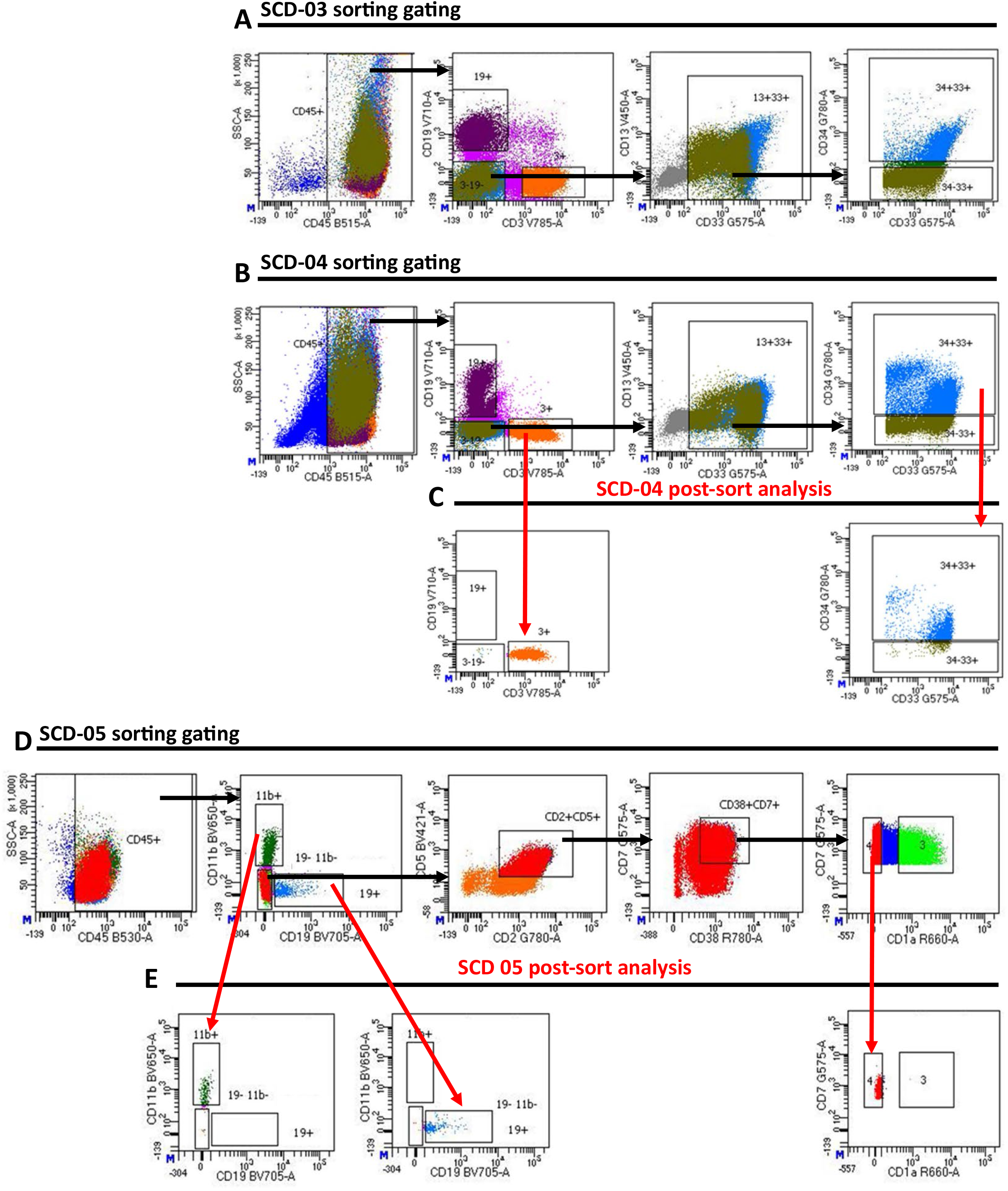
Gating strategies for sorting samples with post-sort analysis. **A)** Gating strategy of SCD-03 PB sample. **B)** Gating strategy of SCD-04 PB sample. **C)** Post-sort analysis of SCD-04 sample showing sorted CD3^+^ T cells (left) and CD3^-^CD19^-^CD13^+^CD33^+^CD34^+^ cells (right). **D)** Gating strategy of SCD-05 BM sample. **E)** Post-sort analysis of SCD-05 sample showing sorted CD11b^+^ myeloid cells (left), CD19^+^ B cells (middle) and, CD19^-^CD11b^-^CD2^+^CD5^+^CD7^+^CD38^+^CD1a^-^ cells (right). Black arrows indicate gating sequence and red arrows indicate post-sort analyses.

## Notes

### Competing Interest Statement

The authors have declared no competing interest.

### Author Declarations

All samples were collected under an NHLBI IRB-approved protocol, clinicaltrials.gov identifiers NCT00977691, NCT00061568, and NCT02105766.

## References

1. Fitzhugh CD, Cordes S, Taylor T, et al. At least 20% donor myeloid chimerism is necessary to reverse the sickle phenotype after allogeneic HSCT. Blood. 2017;130(17):1946–1948.

2. Lawal RA, Mukherjee D, Limerick EM, et al. Increased incidence of hematologic malignancies in SCD after HCT in adults with graft failure and mixed chimerism. Blood. 2022;140(23):2514–2518.

3. Abelson S, Collord G, Ng SWK, et al. Prediction of acute myeloid leukaemia risk in healthy individuals. Nature. 2018;559(7714):400–404.

4. Gondek LP, Sheehan VA, Fitzhugh CD. Clonal Hematopoiesis and the Risk of Hematologic Malignancies after Curative Therapies for Sickle Cell Disease. J Clin Med. 2022;11(11).

5. Mukherjee D, Lawal RA, Fitzhugh CD, Hourigan CS, Dillon LW. TP53 mutation screening for patients at risk of myeloid malignancy. Leukemia. 2024;38(7):1604–1608.

6. Jones RJ, DeBaun MR. Leukemia after gene therapy for sickle cell disease: insertional mutagenesis, busulfan, both, or neither. Blood. 2021;138(11):942–947.

